# Aerosol decontamination and spatial separation using a free-space LED-based UV-C light curtain

**DOI:** 10.1101/2021.12.16.21267937

**Authors:** Andreas Wieser, Jessica Beyerl, Albrecht v. Brunn, Vincent Rieker, Marcus Rieker, Michael Hoelscher, Christoph Haisch

**Affiliations:** Division of Infectious Diseases and Tropical Medicine, University Hospital, Ludwig-Maximilians-University (LMU) Munich, Germany; German Center for Infection Research (DZIF), Partner Site, Munich, Germany; Max-von-Pettenkofer Institute, Ludwig-Maximilians-University Munich, Munich, Germany; Heilbronn University of Applied Sciences/University Heidelberg, Germany; Faculty Informatics and Mathematics, University of Applied Sciences Dresden, Germany; Chair of Analytical Chemistry, Technical University of Munich

**Keywords:** UV-C, UV, LED, light emitting diode, SARS-CoV-2, COVID-19, Coronavirus, *Escherichia coli*, Aerosol, *Staphylococcus aureus*, *MHV*

## Abstract

**Background:** The SARS-CoV-2 pandemic demonstrated the vulnerability of our societies to aerosol transmitted pathogens. With no less than 260mio known cases and > 5mio deaths, SARS-CoV-2 is a global catastrophe leading to human and economic losses unprecedented in recent history. Thus, effective methods to limit the spread of aerosol transmitted pathogens are needed. Universal masking and curfew laws are effective but no permanent solution.

**Methods:** A mass producible LED light source emitting homogeneous parallel UV-C light was used as a “light-barrier” to block the spread of infectious aerosols. In an aerosol test channel, Gram-negative and Gram-positive bacteria as well as coronavirus were nebulized and inactivation rates were determined.

**Findings:** With air speeds of 0.1 m s^-1^ an exposure time of 1 s in the UV-C light is obtained. Reduction in CFU for *E. coli* was >3log10 and for *S. aureus* ∼2.8log10. Plug-forming-units of the murine coronavirus (Mouse Hepatitis Virus, MHV) were reduced by about 3log10.

**Interpretation:** The concept of a UV-C light barrier to ward off infectious aerosols if feasible and possible with a light element as described here. Coupled with sensor based activation/deactivation, such a technology could greatly reduce the transmission rates of aerosol transmitted pathogens while not disturbing natural human behaviour. This is an interesting technology allowing a “new normal” in societies after/with SARS-CoV-2.

## Introduction

SARS-CoV-2 is the causative agent of COVID-19, which has spread around the globe at terrifying speed beginning in spring 2020. Currently, as of December 2021, it is estimated that a total of 270 million subjects have been infected and the death toll is estimated to be above 5.3 million^[1]^. The medical evidence suggests that unlike the other coronavirus diseases that we had seen crossing the species border recently, SARS-CoV-1 and MERS, SARS-CoV-2 can be transmitted very efficiently also by pre-symptomatic or asymptomatic individuals^[2,3]^. Recent evidence also suggests that vaccinations are only providing limited protection against further spread of the disease^[4,5]^. Also with the emergence of the Omicron Variant (B.1.1.529), vaccines seem to have decreased massively in their effectiveness^[6,7]^. Transmission of SARS-CoV-2 occurs via the airborne route, and aerosols are a very important factor. Viable SARS-CoV-2 virus particles could be demonstrated in aerosol particles floating in the air for extended periods of times^[8,9]^. This combination of airborne transmission and pre-/asymptomatic spreaders is extremely problematic. Transmission chains can be sustained under a wide variety of circumstances. Especially in all situations where humans meet in confined spaces, or have to share the same rooms for extended periods of time, commonly used hygienic precautions such as using hand sanitizer are ineffective to prevent the spread of the disease.

As the last resort, universal masking of all people is perceived around the globe now, as the only possible option to suppress the spread of the virus. However, this approach is not always feasible as food consumption or drinking are also routinely performed by subjects in the workplace or private spaces, and this is not compatible with continuously wearing masks. Besides, only few individuals wear the FFP2 masks properly, most often they are leaking resulting in less protection than expected. Last but not least, constantly wearing a mask, especially of FFP2 type, is a nuisance and adherence to the rules to continuously wear masks is thus low in many work- and private settings. With proper use of the masks, adverse health effects are also not completely excluded, especially in populations vulnerable for severe COVID-19 disease due to pre-existing lung damage. Work safety regulations in Europe suggest a maximum duration of 75 min for wearing FFP2 protective equipment at a time, followed by a 30 min break. This is impossible in most work or private environments.

Thus, inactivation of airborne pathogens including SARS-CoV-2 is a desirable goal. This can be obtained by centralised ventilation systems equipped with HEPA filtration and sufficient air flow. Such systems are currently only in operation in laboratory facilities or specialised areas in hospitals, such as intensive care units or operating theatres. They are expensive to maintain and the energy and space consumption to house the respective parts are high. Also most commonly used ventilation systems do not offer sufficient air exchange rates or filtration efficiency and thus may even contribute to the spread rather than decreasing it^[10,11]^. Also, most buildings cannot reasonably be retrofitted with elaborate ventilation systems, other solutions are urgently needed. Most approaches utilise room air filtration using HEPA grade filters, sucking the air through the whole room to remove aerosols. Other systems use air ventilation coupled with UV disinfection in a closed system to re-circulate the air in each room. All these systems however do not hinder the diffusion of infectious aerosols from one person to the other, and indeed might even enhance the transport of aerosols from an infected person to a susceptible person by sucking the aerosols in the respective direction.

Our new approach is using bundled UV-C light to generate a curtain-like invisible barrier within a room to block the dissemination of infectious aerosol and droplets in a room, similar to a transparent wall. The specialised optics used was developed and produced in collaboration with industry partners and will be evaluated along with its efficiency for aerosol decontamination within this manuscript.

## Results

The system consists of the components optics, ventilation, and a safety system for switching off the UV-C source when objects enter the light path. Figure 1 shows a schematic of the system.

**Figure 1:**
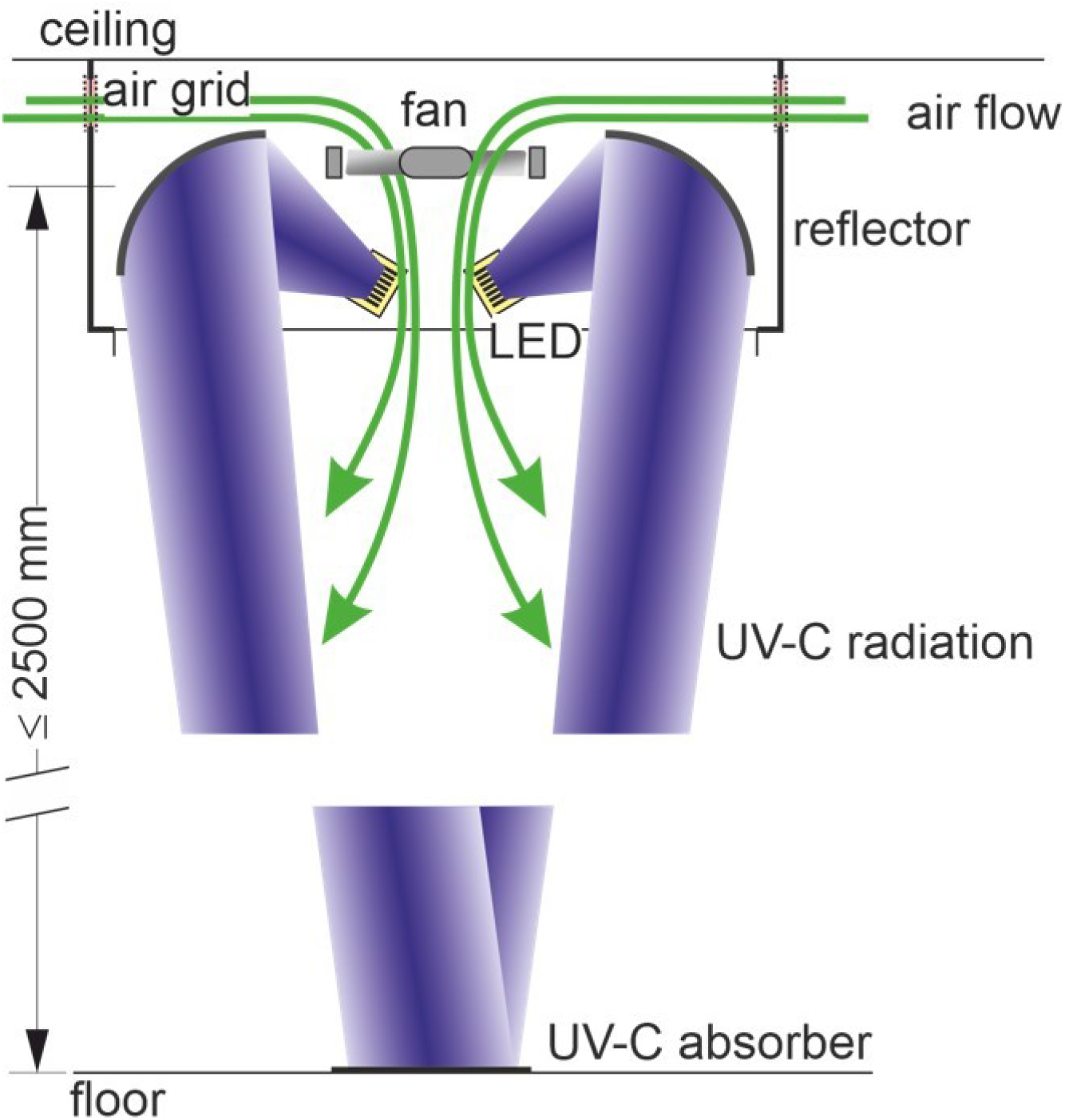
Schematic drawing of the overall system layout. A ceiling mounted housing encloses the mirror based optics and the air duct system.

### Light source

The radiation is generated by multicore LEDs (Luminus, USA). Each LED is projected via a rectangular (62 mm by 62 mm) concave mirror to an area of 100 mm by 200 mm at a distance of z=2,500 mm. Eight of these units are installed in two parallel rows (distance 10 cm) in one curtain module. Before leaving the system, the beams pass through an array of slats which prevent the reflectors from unwanted contact and block stray light. The light emitted by the unit is focussed in one direction, the direction across the curtain, in a distance from the lamp unit of 2,500 mm. A limited divergency in the other direction is required if individual sections are to be switched separately. This feature allows crossing the curtain for human beings as well as for potentially UV-reflecting material without switching off the complete curtain. Economic as well as technical considerations explain the use of reflective optics instead of conventional lens optics.

The UV-C-light beam falls onto a reflector unit to be deflected in vertical direction. The aim is to bundle the radiation density in such a way that a homogeneous radiation volume is created under the arrangement that effectively deactivates viruses as they pass through. At the same time, aerosol is drawn in via the air grids and forced into a gap between the reflectors by transverse fans. The constant flow ensures that the aerosol in the room is sucked into the arrangement and encounters the UV-C radiation as it passes through the reflector unit, or along the UV-C light path to the floor, thus inactivating possible pathogens. In addition, the air flow ensures sufficient cooling of the LEDs.

For the use of UV-C light, it is important to avoid side effects such as the generation of Ozone, if the system is to be used in populated settings, as Ozone is known to be irritative for airways. To generate Ozone, energy rich light with a wavelength of <242 nm is generally considered necessary^[12]^. While many UV-tubular fluorescent lamps emit a broad spectrum including these energy wavelengths, LED based UV-C sources are more narrow band and can be chosen to have a spectrum which will be absorbed efficiently by nucleic acids while not being energy rich enough to produce Ozone. This approach was followed in the light source described here, where LEDs with nominal centre wavelength of 275 nm and a narrow spectrum were used (Luminus, USA). We confirmed that the ozone concentration in the vicinity of the light source was not elevated as expected (data not shown).

### Optical System

With an in-house developed light mapping tool (see Methods chapter), we monitored the optical intensity distribution of the lamp unit in different distances from 500 mm up to a maximum of 2,500 mm, which is considered the usual installation distances of the system above the floor (see Fig. 2). The peak intensity of the curtain at 2,500 nm distance was found to be 2.61 mW cm^-2^, the half width of the curtain at 2500 mm is 53 mm. The intensity maps close to the lamp module clearly illustrate the two parallel rows of LEDs individually, while at distances above 1.5 m, the two rows blend into each other. The two separate rows, however, are not considered a problem, as for the efficiency of the curtain, the integrated dose across the wall exposed to each organism is relevant.

**Figure 2:**
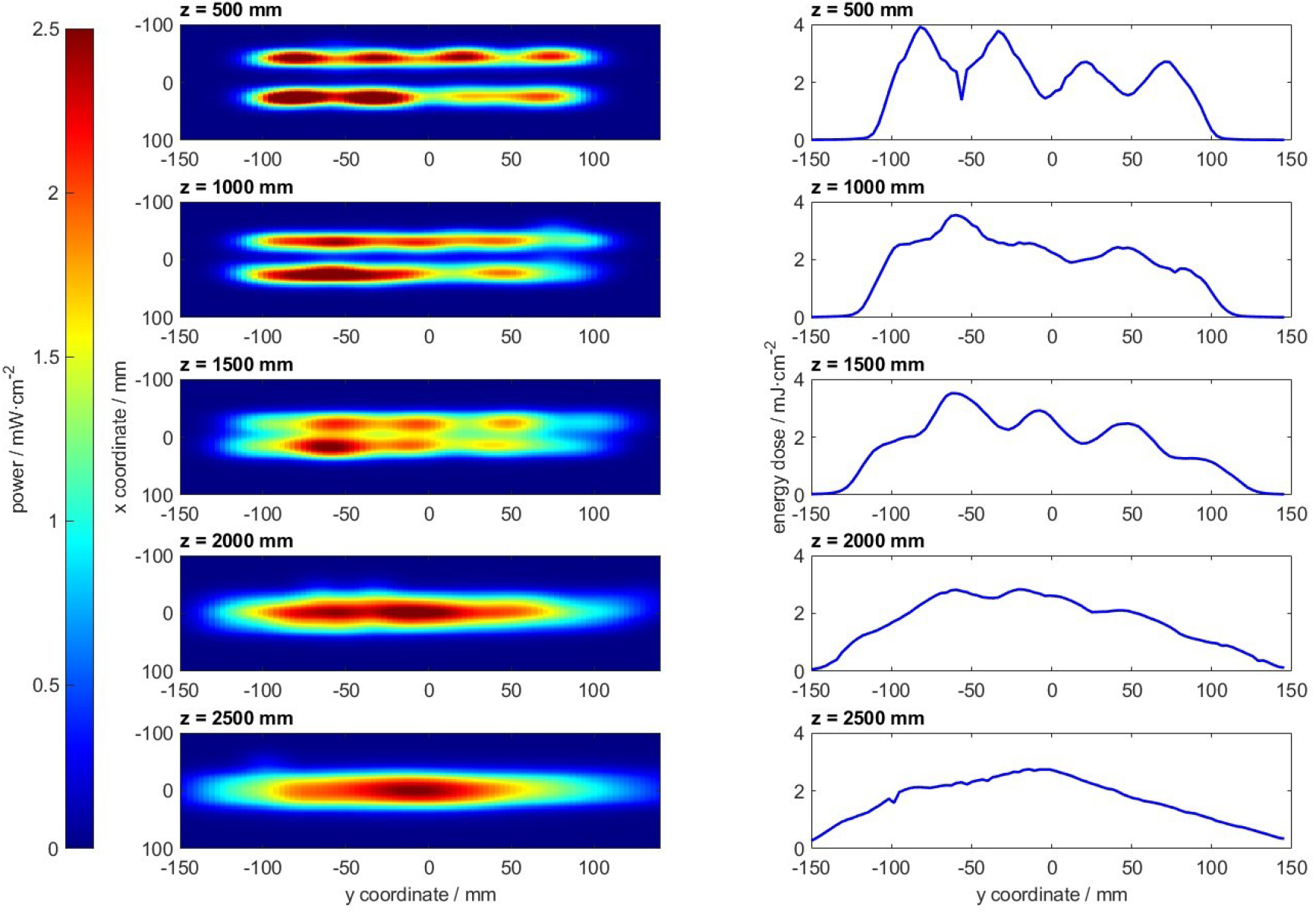
Irradiance map of one optical module in different distances to the source (left) and the corresponding integrated energy dose (right) for a particle passing the light curtain with a speed of 0.1 m s^-1^.

For particles migrating through the wall with a speed of 0.1 m s^-1^, which is the maximum speed to be expected, an integrated UV dose can be calculated by integrating the intensities along the y-axis. These doses are presented in Figure 2 (right side), again for the five different distances from the emitter. It can be appreciated that close to the lamp module, the 4 pairs of LEDs can clearly be distinguished. In consequence, the dose values close to the lamp do not significantly exceed the value of 1.5 mJ considered the minimum for an efficient inactivation of any organism. For COVID-19, threshold values are much lower (∼0.6 mJ).

The optics is adjusted in a way that the superposition of the projections of individual modules, each containing four pairs of LEDs converges at 2,500 mm, to allow operation in rooms with common ceiling height. As we had access to only one module, we simulated a continuous light curtain by mathematically superpositioning the emission of the light distribution of the one module we analysed, with a lateral displacement of 250 mm, which is the length of each module. Figure 3 illustrates the result of a simulated continuous curtain, as well as the same calculation, but simulating the segmented switching of individual modules by the safety shutoff system. Here, the safety shutoff system is engaged at position y=0.

**Figure 3:**
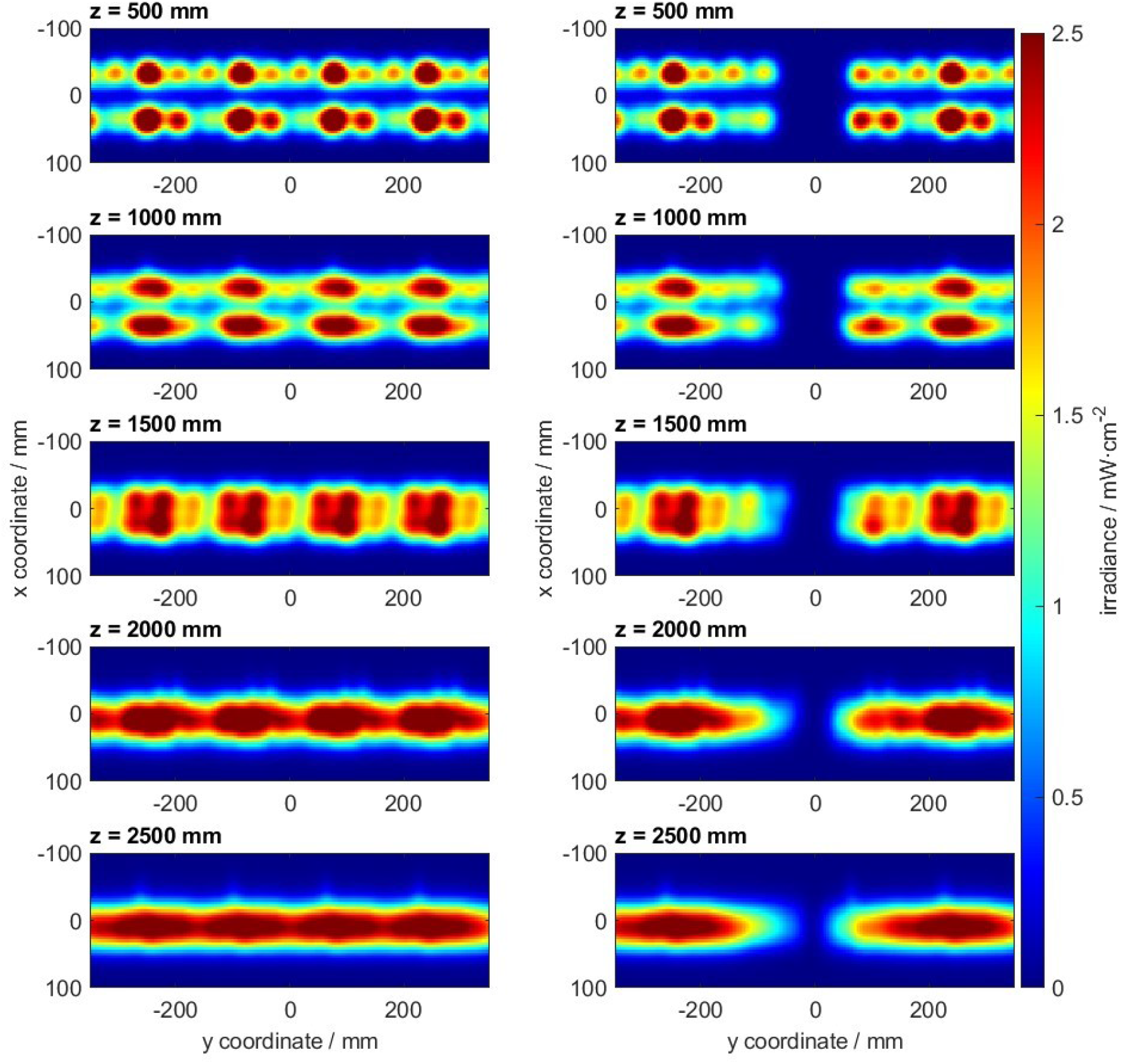
Continuous UV-C curtain simulated mathematically using the measured data of one module (left); the same system with one module in the centre is switched off, simulating a locally engaged safety switch (right).

### Biological Efficiency

Using a UV-C module as described above to illuminate a quartz glass window (100x 100 mm) in an aerosol channel (see supplemental methods for details), biological activity of the system was assessed. In brief, concentrated bacterial or viral solutions respectively were produced and nebulized with a similar size profile encountered in human breath. The aerosols were then directed into a laminar flow channel system with adjusted 45% humidity at 25°C. To retrieve the aerosols after the channel, a gas washing bottle containing 50 ml of phosphate buffered saline (PBS) was used. Serial dilutions were performed and plated in duplicates on Columbia 5% blood agar plates (Becton, Dickinson, Germany) to assess viral load. Virus solutions were inoculated in serial dilutions on L929 cells. Viral plaques and bacterial colonies were manually counted after 10 days or 24 h, respectively.

The experiments resulted in the reduction of culturable bacteria and infectious viruses as shown in table 1. Thereby, in the *E. coli* and the murine coronavirus (Mouse Hepatitis Virus, MHV-A59)^[13]^ group, the infectious load in the aerosol could be reduced to zero in the irradiation condition. Thus, the reduction factor depicted is a conservative estimate and might be higher.

**Table 1:**
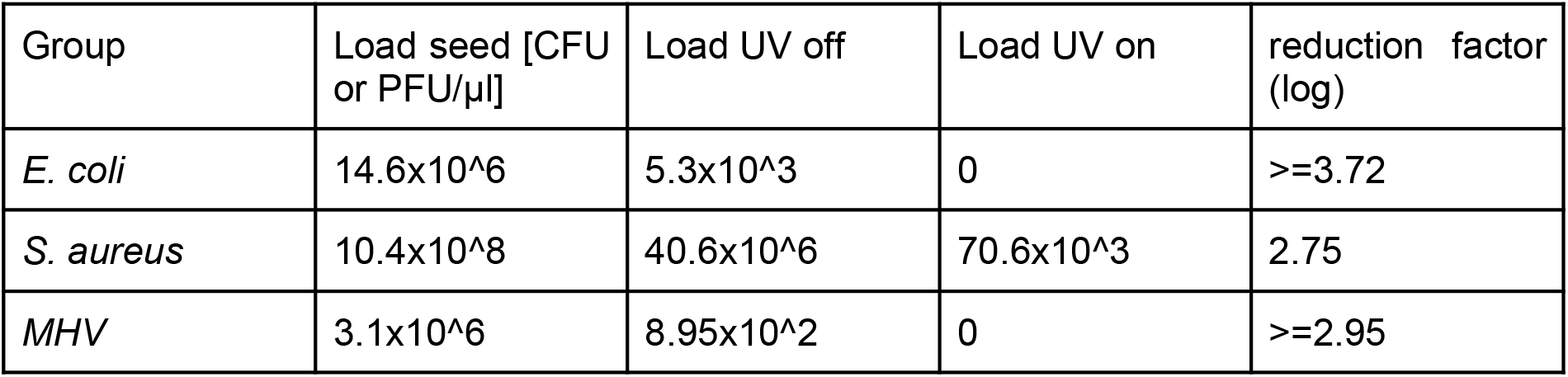
Repetitive load reduction experiments were performed; a representative dataset is shown. Depicted values are averages from three (E.coli and S. aureus) or four (MHV) independent experimental runs. The nebulizer was loaded with the “seed” solution. UV-off and -on conditions were always alternated, starting with the UV-off condition. The reduction factor is calculated as the log10 difference between the UV-exposed and non-exposed group.

| Group | Load seed [CFU or PFU/μl] | Load UV off | Load UV on | reduction factor (log) |
| --- | --- | --- | --- | --- |
| *E. coli* | 14.6x10<sup>6</sup> | 5.3x10<sup>3</sup> | 0 | $\geq 3.72$ |
| *S. aureus* | 10.4x10<sup>8</sup> | 40.6x10<sup>6</sup> | 70.6x10<sup>3</sup> | 2.75 |
| *MHV* | 3.1x10<sup>6</sup> | 8.95x10<sup>2</sup> | 0 | $\geq 2.95$ |

## Discussion

In our setting, we demonstrate a reduction in viable MHV virus in aerosol particles moving through the UV-C light barrier at 0.1 m s^-1^ of almost 3 (2.95) orders of magnitude (99.9%). The UV-C energy transfer that can be assumed is in the range between 1.5 mJ cm^-2^ and 4 mJ cm^-2^ (figure 2), as a distance of 500 mm was used in the flow chamber setup. At this distance, the most unfavourable, and inhomogenous beam pattern was observed in the system. Coronavirus inactivation was assessed in different studies with different virus strains including common cold coronavirus and SARS-CoV-2. However, all studies show similar outcomes^[14-19]^. Other studies measured higher doses of UV-C radiation needed^[20]^, which is not surprising as this model uses viral solutions in a petri dish filled with water based buffer solution, resulting in absorption of UV-C light due to the several millimetre thick water layer, an effect otherwise not observed in aerosols of only few micrometres in diameter. Bacteria are generally more difficult to inactivate with UV-C radiation also because they possess potent repair mechanisms for their genome. As there is no generally accepted protocol to assess the UV-C doses needed to inactivate different bacteria in aerosols, data on the necessary dose for inactivation varies quite drastically, even for *E. coli* K-12 one of the most used model organisms. For example *Malayeri et al*. lists the UV-C dose needed for 1 log reduction in K-12 *E. coli* as between 1.1 and 7.8 mJ cm^-2^, derived from multiple sources^[21]^. Generally again, such data is not available for aerosolized bacteria which will most likely be more vulnerable given that less water is surrounding and protecting them by absorbing UV-light. Thus, we performed inactivation experiments for *E. coli* K-12 as a gram-negative model organism, as well as *S. aureus* a more UV-resistant, but relevant human pathogen. For both we could demonstrate significant load reductions of >99.9 and 99.9% respectively. Under the chosen conditions of 0.1 m s^-1^ airspeed, the UV-C intensity will most likely be insufficient to safely inactivate spores of *Aspergillus spp*. or *Bacillus spp*. as those are described to generally require doses >20 mJ cm^-2^ to be inactivated by at least one order of magnitude^[21]^.

Protection against aerosol transmitted pathogens is generally difficult. The most effective way of protection are respiratory masks with a very tight fit without any leaks. If using the proper filtration materials, almost complete protection can be achieved^[22,23]^. Thereby, the materials filter larger particles with higher efficiency than smaller particles around 0.3 µm^[24]^. The biggest problem in real use however, is the improper fit resulting in leakage of air not passing the filter. For aerosol particles of small diameters, surgical masks show very high leaks of 70% or even more, while unadjusted FFP2 masks also have leaks of 50% and higher. Even after adjustment, FFP2 masks will often have 15% or more leak^[22]^, reducing the overall efficiency of the mask dramatically as leak air is taken in without any filtration whatsoever. It is also commonly observed that many subjects will wear the masks not as tight as a trained professional would, or even adjust the mask to maximise leak in order to breath with less resistance and more comfortably. Despite these low real filtering efficiencies due to leaks, mask wearing is associated with protection against SARS-CoV-2 infection^[25,26]^.

Alternative measures for protection against SARS-CoV-2 transmission through the air is the use of room air filtration systems. While evidence of transmission reduction with the use of air filters is anecdotal at best, it has been solidified that such systems can remove virus-containing particles from the air^[27,28]^. Still, it is quite clear, that such systems can only prevent the accumulation of infectious aerosols in closed rooms while direct transmission between persons also via aerosols is still very possible, and could even be enhanced when the airflow generated by the filter is drawing infectious aerosol towards s susceptible person before filtering it. Thus, wearing of masks is still advised as a personal protection measure^[27]^.

The use of solid transparent glass or plastic barriers is providing protection against ballistic droplets, however aerosols will float around such barriers depending on the air movement, if the barrier is not sealed to the ceilings and walls. Nevertheless, experimental studies have found significant reductions of aerosol particles if the barrier is large enough and correctly positioned^[29]^. Such hard mechanical barriers however restrict the movement and are often not compatible with many activity areas. Still, some limited protection against direct transmission can be assumed.

All this demonstrates that protection against SARS-CoV-2 infection can be achieved by multiple different interventions. Even the best widely accepted measure, the use of FFP2 masks, reduces the uptake of infectious aerosols often only less than 90% due to leaks^[22]^. Also these leaks will be considerably bigger during sneezing or coughing due to the backpressure of the mask^[23]^. For interventions such as air filters or solid barriers it is difficult to assess the reduction in infectious dose for individual encounters between infected and uninfected subjects, still both can be considered to be less efficient than masking.

With a light barrier system as described in this manuscript, a rather homogeneous light curtain that is permanently active and open to the rest of the air volume in the respective space is produced. There is no controlled exposure time of aerosols or droplets within the irradiation field. Rather, the UV-C dose is directly proportional to the duration of stay inside the irradiated air volume. So, the dose depends on the speed of the individual aerosols moving across the UV-C-illuminated volume, i.e. the local air speed, and on the width of the light curtain. To assess realistic conditions, an airspeed of 0.1 m s^-1^ was chosen in this study. This is the maximum speed measured in 1 m distance from a human mouth after sneezing, which is the fastest air speed the human body generates at a distance of about 1 m^[30]^. Lower air movement speed will obviously be the condition encountered in most rooms, resulting in even higher rates of inactivation, not assessed in this study.

As evident e.g. in figure 2, homogeneity of the light curtain is still an issue in our prototype system, some LED offer less output then others. This is due to variation in the used pre-series LEDs. The commercial product, which is planned to be available beginning of 2022 will, according to the producer, feature very low standard deviation (as common for commercial LEDs) and even slightly more optical output power than the ones used in this study. The very fast development of high-power UV-C LEDs observed in the last years and even months will be most beneficial for the UV light curtain.

If a UV-C light barrier is mounted in a way that air cannot pass around it, e.g. by mounting it between solid walls, aerosol viral load reductions higher than those observed with FFP2 masks are achievable as we have demonstrated. Besides, there are indications for aerosol transmissible viruses such as SARS-CoV-2, that they can also be taken up by impaction of infectious droplets on the conjunctiva^[31,32]^. To protect against this route of infection, masks are insufficient and specialised goggles would be additionally required. Obviously, masks as well as goggles also obstruct the voluntary or involuntary contact between potentially contaminated fingers and the eyes, nostrils or the mouth, an effect otherwise not achieved by any other commonly used measure. On the other hand, a light barrier would inactivate infectious aerosols before they reach the eyes of another person, thus providing additional protection, a combination not achieved with any other system currently used. Also in certain areas, such as during consumption of food, wearing a mask is impossible. Further, a light barrier is a technical system that, if functional, has less variation in its efficiency than e.g. masking with a wide variation of performance due to user errors or anatomy of the wearer.

Exposure to UV-C radiation can have a negative impact on health, especially when it affects the skin or the eyes. The health risk is separated into immediate damage which is in most cases reversible and any long-term damage which is difficult to link to the actual exposure event. The extent of damage to tissue is determined by the length of time (exposure duration) for which a tissue is exposed to radiation and the irradiance of the UV source. In both the USA and Europe, the maximum daily dose for wavelengths from 180 to 400 nm is 30 J m^-2^ for an exposure time of 8 hours, which corresponds to one work day (Directive 2006/25/EC, DIN EN 62471 (VDE 0837-471):2009-02, DIN EN 14255, ISO 15858:2016, American Conference of Governmental Industrial Hygienists (ACGIH). Threshold Limit Values (TLVs) for occupational exposure to UV). This means that a potential use of an open UV-C light barrier requires a safety mechanism, securely shutting down the system when a body part or reflective object enters the UV-C irradiated area. In order to maintain maximum efficiency of aerosol inactivation, only the affected parts of the light barrier within a room shut down. Also, stray light has to be minimised to reduce any hazardous potential.

A further potential source of stray radiation would be the floor underneath the system. Thus, a high strength absorber layer is placed. It is less than a millimeter thick and is attached (glued) directly to the floor. The absorber material was chosen so that it can withstand long irradiation times without showing signs of dissolution. In addition, the safety device detects objects near the beam path and switches off the irradiation to avoid reflections.

## Data Availability

Raw data will be made available upon reasonable request to fellow colleagues in the field.

## Acknowledgements

The company Smart United for developmental support and provision of materials and support. The work was supported by grants of the “Bundesministerium fuer Bildung und Forschung’’ of the German Government (RAPID 01Kl1723C/01KI2006C) to AvB (Ludwig-Maximilians-University Munich), and the German Center for Infection Research (DZIF, partner site Munich, project TTU EI 01.806 to AvB).

## Author contribution

AW, MR and CH designed the protocol, JB, AvB, VR, CH and AW performed laboratory work, AW, AvB, JB, CH, MR, MH analysed the data and drafted the manuscript.

## References

1 World Health Organization, WHO. Coronvirus (COVID-19) Dashboard, December 2021 https://covid19.who.int/

2 Rothe, C. et al. Transmission of 2019-nCoV Infection from an Asymptomatic Contact in Germany. N Engl J Med 382, 970–971, doi:10.1056/NEJMc2001468 (2020).

3 Muller, C. P. Do asymptomatic carriers of SARS-COV-2 transmit the virus? Lancet Reg Health Eur 4, 100082, doi:10.1016/j.lanepe.2021.100082 (2021).

4 Eyre, D. W. et al. The impact of SARS-CoV-2 vaccination on Alpha & Delta variant transmission. medRxiv, 2021.2009.2028.21264260, doi:10.1101/2021.09.28.21264260 (2021).

5 Kroidl, I. et al. Vaccine breakthrough infection and onward transmission of SARS-CoV-2 Beta (B.1.351) variant, Bavaria, Germany, February to March 2021. Euro Surveill 26, doi:10.2807/1560-7917.Es.2021.26.30.2100673 (2021).

6 Holgersen, E. M. et al. Transcriptome-Wide Off-Target Effects of Steric-Blocking Oligonucleotides. Nucleic Acid Ther 31, 392–403, doi:10.1089/nat.2020.0921 (2021).

7 Cele, S. et al. SARS-CoV-2 Omicron has extensive but incomplete escape of Pfizer BNT162b2 elicited neutralization and requires ACE2 for infection. medRxiv, 2021.2012.2008.21267417, doi:10.1101/2021.12.08.21267417 (2021).

8 Nissen, K. et al. Long-distance airborne dispersal of SARS-CoV-2 in COVID-19 wards. Sci Rep 10, 19589, doi:10.1038/s41598-020-76442-2 (2020).

9 van Doremalen, N. et al. Aerosol and Surface Stability of SARS-CoV-2 as Compared with SARS-CoV-1. N Engl J Med 382, 1564–1567, doi:10.1056/NEJMc2004973 (2020).

10 Pease, L. F. et al. Investigation of potential aerosol transmission and infectivity of SARS-CoV-2 through central ventilation systems. Build Environ 197, 107633, doi:10.1016/j.buildenv.2021.107633 (2021).

11 Almilaji, O. Air Recirculation Role in the Spread of COVID-19 Onboard the Diamond Princess Cruise Ship during a Quarantine Period. Aerosol and Air Quality Research 21, 200495, doi:10.4209/aaqr.200495 (2021).

12 Claus, H. Ozone Generation by Ultraviolet Lamps(†). Photochem Photobiol 97, 471–476, doi:10.1111/php.13391 (2021).

13 Züst, R. et al. Coronavirus non-structural protein 1 is a major pathogenicity factor: implications for the rational design of coronavirus vaccines. PLoS Pathog 3, e109, doi:10.1371/journal.ppat.0030109 (2007).

14 Buonanno, M., Welch, D., Shuryak, I. & Brenner, D. J. Far-UVC light (222 nm) efficiently and safely inactivates airborne human coronaviruses. Sci Rep 10, 10285, doi:10.1038/s41598-020-67211-2 (2020).

15 Robinson, R. T., Mahfooz, N., Rosas-Mejia, O., Liu, Y. & Hull, N. M. SARS-CoV-2 disinfection in aqueous solution by UV<sub>222</sub> from a krypton chlorine excilamp. medRxiv, 2021.2002.2019.21252101, doi:10.1101/2021.02.19.21252101 (2021).

16 Storm, N. et al. Rapid and complete inactivation of SARS-CoV-2 by ultraviolet-C irradiation. Sci Rep 10, 22421, doi:10.1038/s41598-020-79600-8 (2020).

17 Sabino, C. P. et al. UV-C (254 nm) lethal doses for SARS-CoV-2. Photodiagnosis Photodyn Ther 32, 101995, doi:10.1016/j.pdpdt.2020.101995 (2020).

18 Heilingloh, C. S. et al. Susceptibility of SARS-CoV-2 to UV irradiation. Am J Infect Control 48, 1273–1275, doi:10.1016/j.ajic.2020.07.031 (2020).

19 Heßling, M., Hönes, K., Vatter, P. & Lingenfelder, C. Ultraviolet irradiation doses for coronavirus inactivation - review and analysis of coronavirus photoinactivation studies. GMS Hyg Infect Control 15, Doc08, doi:10.3205/dgkh000343 (2020).

20 Ma, B., Gundy, P. M., Gerba, C. P., Sobsey, M. D. & Linden, K. G. UV Inactivation of SARS-CoV-2 across the UVC Spectrum: KrCl* Excimer, Mercury-Vapor, and Light-Emitting-Diode (LED) Sources. Appl Environ Microbiol 87, e0153221, doi:10.1128/aem.01532-21 (2021).

21 Malayeri, A., Mohseni, M. & Cairns, B. Fluence (UV Dose) Required to Achieve Incremental Log Inactivation of Bacteria, Protozoa, Viruses and Algae. IUVA News 18, 4–6 (2016).

22 Bagheri, G., Thiede, B., Hejazi, B., Schlenczek, O. & Bodenschatz, E. An upper bound on one-to-one exposure to infectious human respiratory particles. Proceedings of the National Academy of Sciences 118, e2110117118, doi:10.1073/pnas.2110117118 (2021).

23 Schmitt, J. & Wang, J. Quantitative modeling of the impact of facemasks and associated leakage on the airborne transmission of SARS-CoV-2. Scientific Reports 11, 19403, doi:10.1038/s41598-021-98895-9 (2021).

24 Tcharkhtchi, A. et al. An overview of filtration efficiency through the masks: Mechanisms of the aerosols penetration. Bioact Mater 6, 106–122, doi:10.1016/j.bioactmat.2020.08.002 (2021).

25 Chu, D. K. et al. Physical distancing, face masks, and eye protection to prevent person-to-person transmission of SARS-CoV-2 and COVID-19: a systematic review and meta-analysis. The Lancet 395, 1973–1987, doi:10.1016/S0140-6736(20)31142-9 (2020).

26 Cheng, Y. et al. Face masks effectively limit the probability of SARS-CoV-2 transmission. Science, doi:10.1126/science.abg6296 (2021).

27 Kähler, C., Fuchs, T., Mutsch, B. & Hain, R. Schulunterricht während der SARS-CoV-2 Pandemie -Welches Konzept ist sicher, realisierbar und ökologisch vertretbar?, (2020).

28 Conway Morris, A. et al. The removal of airborne SARS-CoV-2 and other microbial bioaerosols by air filtration on COVID-19 surge units. Clin Infect Dis, doi:10.1093/cid/ciab933 (2021).

29 Bartels, J., Estill, C. F., Chen, I.-C. & Neu, D. Laboratory Study of Physical Barrier Efficiency for Worker Protection against SARS-CoV-2 while Standing or Sitting. medRxiv, 2021.2007.2026.21261146, doi:10.1101/2021.07.26.21261146 (2021).

30 Ivanov, M. Exhaled air speed measurements of respiratory air flow, generated by ten different human subjects, under uncontrolled conditions. E3S Web of Conferences 111, 02074, doi:10.1051/e3sconf/201911102074 (2019).

31 Szczęśniak, M. & Brydak-Godowska, J. SARS-CoV-2 and the Eyes: A Review of the Literature on Transmission, Detection, and Ocular Manifestations. Med Sci Monit 27, e931863, doi:10.12659/msm.931863 (2021).

32 Coroneo, M. T. & Collignon, P. J. SARS-CoV-2: eye protection might be the missing key. Lancet Microbe 2, e173–e174, doi:10.1016/s2666-5247(21)00040-9 (2021).

