## Supplemental Methods for "Aerosol decontamination and spatial separation using a free-space LED-based UV-C light curtain"

**Affiliations:**

Elisabeth-Winterhalter-Weg 6, 81377 Munich, Germany

**KeyWords:**

UV-C, UV, LED, light emitting diode, SARS-CoV-2, COVID-19, Coronavirus, *Escherichia coli*, Aerosol, *Staphylococcus aureus, MHV*

**Aerosol decontamination and spatial separation using a free-space LED-based UV-C light curtain**

Andreas Wieser^1,2,3*^, Jessica Beyerl^1^, Albrecht v. Brunn^2,3^, Vincent Rieker^4^, Marcus Rieker^5^ Michael Hoelscher^1,2^, Christoph Haisch^6*^

### Online supplement

Optical Mapping

Optical mapping was performed by an in-house made device. An UV-3726 irradiance detector, calibrated for a spectral range from 260 nm to 290 nm in intensity values (mW cm^-2^), in combination with an optical power meter (X1-5 Optometer, Gigahertz-Optik, Germany) was installed on an xy stage with a scanning range of 630 nm (x direc.) to 450 mm (y direc.). The optical probe head was scanned stepwise (3.2 mm by 4 mm) over the complete range and the optical power was recorded for each individual data point by means of a in-house programmed software tool. Recording of a complete map takes 33 minutes. The detector provides a linear detection range from 0.002 µW∙cm^-2^ to 1,000 mW∙cm^-2^.

Biological Efficiency

For all efficiency tests, bioaerosol had to be generated by nebulization of dispersions of the respective organism in water. In brief, the gram negative bacterium *Escherichia coli* (*E. coli*) strain K-12 and the gram positive bacterium *Staphylococcus aureus* (*S. aureus*) strain MP2235 (isolate) were grown in LB (Luria-Bertani) media overnight to reach a high optical density OD600 > 3.5. The bacterial culture was harvested by centrifugation and washed 2 times with PBS. Bacterial density was determined by plating serial dilutions in duplicates on 5% sheep blood Columbia agar plates (Becton-Dickinson, Heidelberg, Germany).

To assess the effectiveness against Coronavirus, the Mouse Hepatitis Virus strain MHV-A59^[13]^ was reared on L929 cells. High-titer virus stock was produced by trypsinizing two 10 cm dishes of L929 cells. Trypsin was inactivated by adding DMEM containing 10% FCS and pelleting cells at 1000 rpm for 5 min. Cells were resuspended in 4 ml DMEM without FCS in a 15 ml Falcon tube. Virus was added and cells were gently agitated for 2 h at RT. Infected cells were then distributed to four 175 cm² flasks in 20 ml DMEM/10% FCS each, and cultivated for four days at 33°C/5% CO_2_. After four days supernatants were collected, pooled and pre-cleared at 3,000 rcf for 10 min. Virus-containing supernatants were concentrated by Vivaspin 10K at 3,000 rcf for 30 min at RT. Paque titration revealed a virus titer of 10^9 pfu ml^-1^.

To determine the biological effectiveness of the system, an aerosol channel was built (see schematic drawing Supplemental Figure 2). As a safety precaution, the flow channel was constructed to be placed inside a class II safety cabinet in a BSL 2 laboratory. The channel features a rectangular cross section with a width of 100 mm and 50 mm depth. Inside the channel, a laminar flow is maintained. The laminar flow field inside the channel is covered from two sides by fused silica windows (10 cm by 10 cm), allowing the UV light to pass orthogonally through the channel in a way that all particles flowing through the channel are exposed to a homogeneous optical field. Although fused silica is transparent to UV-C light, a reduction of the output in the model of about 10% is expected due to reflection and absorption. These effects do not occur in the final applications.

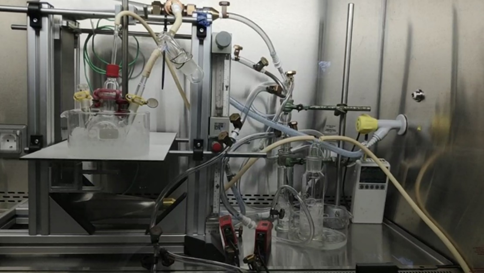

**Supplemental Figure 1:** *Picture of the experimental setup for inactivation of infectious aerosol.*

For the experiment, an airflow velocity perpendicular to the lamp axis (see Supplemental Figure 2) of 0.1 m s^-1^ was selected.

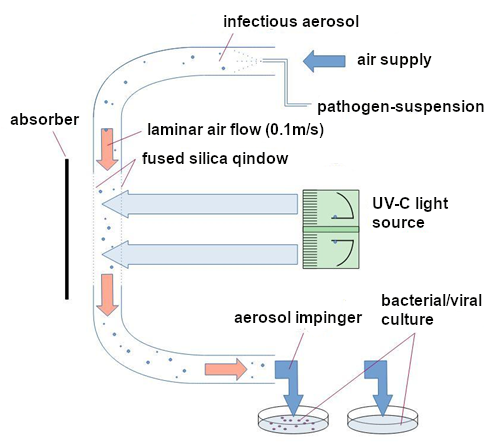

**Supplemental Figure 2:** *Experimental setup for inactivation of infectious aerosol.*

Moisture and temperature controlled supply air (45% at 25°C) is enriched with bacteria/viruses by cross-flow nebulization. An aerosol is formed from the constantly ice-cooled suspension with a size distribution similar to the aerosol in human breath. Through a UV-transmissive window, the aerosol in the flow channel is irradiated with UV-C light. To avoid reflection artefacts, the UV-C light is absorbed behind the channel. The flow velocity in the channel cross-section (100x50 mm^2^, 30 l min^-1^ this corresponds to 0.1 m∙s^-1^). After passing through the UV-C light, the aerosol is collected over a 25 minutes time period in an ice-cooled gas washing bottle. In every experiment, multiple runs were performed with alternating conditions. “UV switched off” was always used as the first condition and in subsequent runs the UV on/off were alternated. After the experimental series, the whole channel was decontaminated internally by the use of a hot air gun, heating the system to > 180°C for > 5 min.

To retrieve viable bacteria collected from the aerosols after the channel, the 50ml of phosphate buffered saline (PBS) in the gas washing bottle were concentrated by centrifugation (10,000 rpm, 4°C, 10 min). Thereafter, the bacterial pellet was resuspended in 100 µl of ice cold PBS. 50 µl of this were plated directly (once), the other 50 µl were diluted 1:10 in serial dilutions down to x10^-9^. For each dilution step 50 µl were plated in duplicates on 5% Columbia sheep blood agar (Becton Dickinson, Germany). Colonies were counted manually after 24 h of incubation at 37°C under aeration. Plates with less than three or more than 1,000 colonies were no longer counted.

For virus, concentration was performed using Sartoris Vivaspin centrifugal concentrator with a MWCO of 30 kDa at 6,000xg for 20 min at 4°C. The virus particles were resuspended from the wet filter membrane with 200 µl of DMEM/2% FCS resulting in a total sample volume of 250 µl. To determine MHV-A59 virus titers after concentration, L929 cells were split onto 24 well cell culture plates. At about 70% confluence, cells were infected with 200 µl virus dilution (n=2) ranging from 10^0 to 10^-9 for one hour at 37°C/5% CO_2_, followed by spin inoculation at 225 rcf for 20 min at 22°C in a Sigma centrifuge 6K15 using swing-out rotor 1115/13229, again followed by incubation at 37°C/5% CO_2_ for 45 min. Infection medium was removed, cells were overlaid with 1.5% carboxymethylcellulose in DMEM/1% FCS and incubated for 10 days at 33°C. Then cells were fixed at 5% paraformaldehyde for 30 min at RT. After two PBS washes, cells were stained with 1% *Crystal Violet* dissolved in 20% ethanol for one hour and washed several times with water until plaques could be discriminated. Plaques were counted manually.
