## Supplementary figures and images for "Aerosol decontamination and spatial separation using a free-space LED-based UV-C light curtain"

### Supplemental Figure1

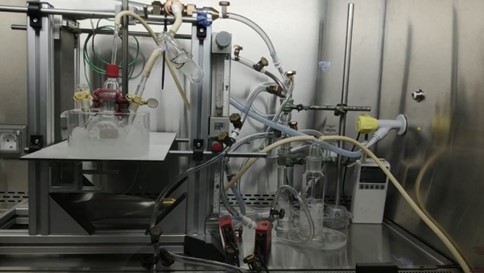

### Supplemental Figure 2

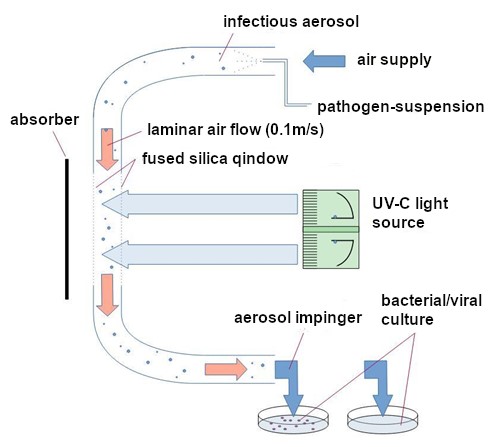
